# What strategies are used to select patients for direct admission under acute medicine services? A systematic review of the literature

**DOI:** 10.1101/2024.09.10.24313268

**Authors:** Samuel Evans, Catherine Atkin, Austin Hunt, Georgina Ball, Charlotte Cassidy, Alexander Costley-White, Louise Toy, Rebecca Wilding, Elizabeth Sapey

## Abstract

**Background:** Pressures on hospital emergency care services have led to increasing interest in new models of acute care provision. One such model is a medical emergency department where medical patients are triaged directly to acute internal medicine, without assessment by emergency medicine. The evidence for this model of care is unclear.

**Design:** Systematic review.

**Methods:** Studies included direct referral pathways to acute internal medicine. The protocol was registered prospectively (Prospero: CRD42023495786). Databases searched included MEDLINE (Ovid), The Cochrane Central Register of Controlled Trials (CENTRAL), MEDLINE in process, Web of Science, CINAHL, and Embase. Studies had no time or language restrictions. Studies were selected based on inclusion and exclusion criteria, assessed by at least two independent researchers. ROBINS I risk of bias assessment was applied to the selected studies and a narrative synthesis was performed.

**Results:** From 4405 abstracts, 89 full text articles were screened and 4 were selected for data extraction. Two studies assessed tools to predict the need for a medical admission and two studies assessed the impact of direct referral pathways to medicine. Risk of bias was mixed, and studies were heterogeneous. However, the studies reported a good ability to appropriately select patients for direct referral to medicine and a reduced length of time to medical assessment. There were no differences in other outcomes such as mortality or overall length of stay.

**Discussion:** The current evidence to support direct admission to medicine, effectively a medical ED, is limited with studies being heterogeneous and of varying quality. Models for patient selection varied, but there was evidence to support accurate, early identification of medical patients and of reduced delays in medical assessment and care.

**Conclusion:** Given these positive early signs of benefit, more studies are needed to design and evaluate care models such as medical EDs.

**Registration:** Prospero Registration Number: CRD42023495786.

**What is already known on this topic:** Direct admission pathways to acute medicine services are used in some centres in the UK with significant variation in how this pathway is provided.

**What this study adds:** This systematic review is the first comprehensive synthesis of published research on direct admission pathways to internal medicine services. The limited number of studies were heterogenous and of variable quality. Different models for patient selection were included but were assessed, studies demonstrated the ability to identify patients likely to require medical admission, and a reduction in the time to medical admission. More studies are needed to assess how to structure and operationalize a direct admission pathway in the United Kingdom and internationally.

**How this study might affect research, practice or policy:** Our study highlights the need for further research to help develop optimal pathways to enable patients with acute medical conditions requiring treatment to be reviewed by acute medical teams as soon as possible after presentation, to improve patient care in the context of growing demand for these services.

**Strengths and Limitations:**

- This is the first systematic review of direct admission pathways to medicine.
- The systematic review was conducted using standardised methodology with the protocol prospectively registered on an open access database. There were no date or language restrictions applied.
- The main limitation of the systematic review is the limited number and quality of studies available for inclusion.

## Introduction

Urgent and emergency care (UEC) services face unprecedented challenges and demand with approximately 2.5million emergency internal medicine admissions every year in England^[1]^. Medical emergencies are the most common reason for attendance to emergency departments and are predominantly referred to acute medical services^[2]^ with evidence showing that early assessment of patients by acute internal medical (AIM) services improves patient outcomes^[4]^.

Traditional routes to medicine services involve initial triage and assessment by emergency medicine teams, then often a senior clinical review by emergency medicine, followed by subsequent referral to acute internal medicine, an assessment by a tier one clinical decision maker in acute medicine, followed by an assessment by a senior acute internal medicine healthcare professional.

The Society for Acute Medicine has published guidelines on best clinical practices stating that a review by a senior acute medicine clinician should occur within 6 – 14 hours from referral to acute medicine depending on time, with a 6-hour target during the day and a 14-hour target out of hours. At present, only 41% of patients referred to acute medicine are reviewed within this timeframe^[3,6]^.

With the changing demographics of our population, current trajectories of acute medicine demand describe increases year on year, outstripping the ability of UEC services to provide care, leading to overcrowding in emergency departments and long delays for patients and poorer outcomes^[4]^. The performance of front door services is widely expected to decrease unless an intervention is made. Innovative approaches to streamline patient care and improve patient experience and outcomes are needed^[4]^.

The Getting it Right First-Time national report outlines the need to move to an emergency department streaming model of patient care where following an initial triage the patient can be moved directly to a specialist area for assessment^[5]^. For most patients, this involves transfer to AIM services including the acute medical unit (AMU), same day emergency care (SDEC) unit or other such AIM-delivered areas. The model of triage to AIM has been described as a “Medical Emergency Department” or “Medical Referral Unit,” with patients who would benefit from care from AIM teams moving directly to an AIM unit after initial triage, without review by Emergency medicine clinical teams.

Previous systematic reviews demonstrated that there is a lack of evidence to support how best to deliver acute medical care in the UK and the Society for Acute Medicine annual benchmarking audit has identified significant heterogeneity in how services are run^[6, 7]^. With growing interest in direct admissions pathways in acute medicine it is important to review and assess the evidence base that exists prior to the development of any streaming pathways.

This systematic review specifically aimed to answer the questions of which direct admission pathways have been developed, and what the evidence was for their effectiveness, including safety and impact on patient outcomes compared to the traditional pathway through the Emergency Department.

The systematic review has two main objectives, 1. To identify the approaches used to identify and select patients directly admitted to acute medicine services from the emergency department. 2. To compare the safety and operational efficiencies of such models compared to usual care (defined as being where patients are reviewed by ED clinicians and then referred to onwards to AIM teams).

## Methods

This review was undertaken and reported in accordance with PRISMA guidelines, conducted using validated methodology^[8]^ and the protocol was prospectively registered on an open access registry prior to data collection (PROSPERO CRD42023495786 https://www.crd.york.ac.uk/prospero/).

### Search strategy

A total of 6 online databases were searched: MEDLINE (Ovid), The Cochrane Central Register of Controlled Trials (CENTRAL), MEDLINE in process, Web of Science, CINAHL, Embase. The search was undertaken focusing on all reported studies with an end date of 24th of January 2024. There was no limitation placed to language. The search method was conducted using a PICO format seen in Table 1.

**Table 1:** The PICO structure for the systematic review.

|  |  |
| --- | --- |
| Population | Adult patients requiring hospital acute medical services, attending the emergency department, via self-referral, ambulance conveyance or primary care referral. |
| Intervention | Direct medical admission to acute medical services |
| Comparison | Adult patients who have been review by Emergency Department teams and requiring acute medical services |
| Outcomes: | *Primary outcomes:*<br>Time until review by a medical senior clinical decision maker<br>*Secondary outcomes:*<br>Mortality (in hospital and 30 day)<br>Length of stay<br>Readmission rates in 7 and 30 days<br>Time until review by an acute medicine specialist<br>Time to review by a tier one clinical decision maker.<br>Utilisation of Same Day Emergency Care pathways (SDEC)<br>Comparison of any inclusion or exclusion criteria used for direct admission pathways. |

The search conducted was broad to reflect the wide nomenclature of acute medicine services internationally and was mapped to MeSH terms found in each database where possible. Boolean operators OR were used between search terms in each category and the operator AND between each of the categories. A full list of the search terms used in each database can be found in the supplementary data of the online supplement.

References identified were imported into the Covidence software package (Covidence, Australia). Duplicates were removed, and remaining titles and abstracts were screened against eligibility criteria by two reviewers working independently. Where there was disagreement, and if a consensus decision was not possible, final adjudication was provided by an independent senior team member. Full text review was then undertaken against the same criteria and using the same process. The eligibility criteria are listed in Table 2.

**Table 2:**
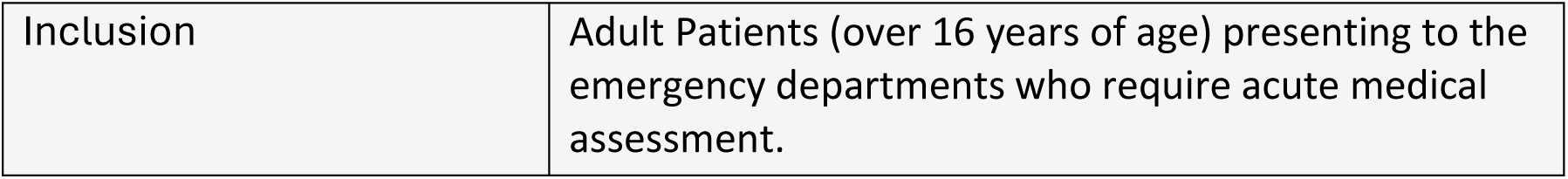
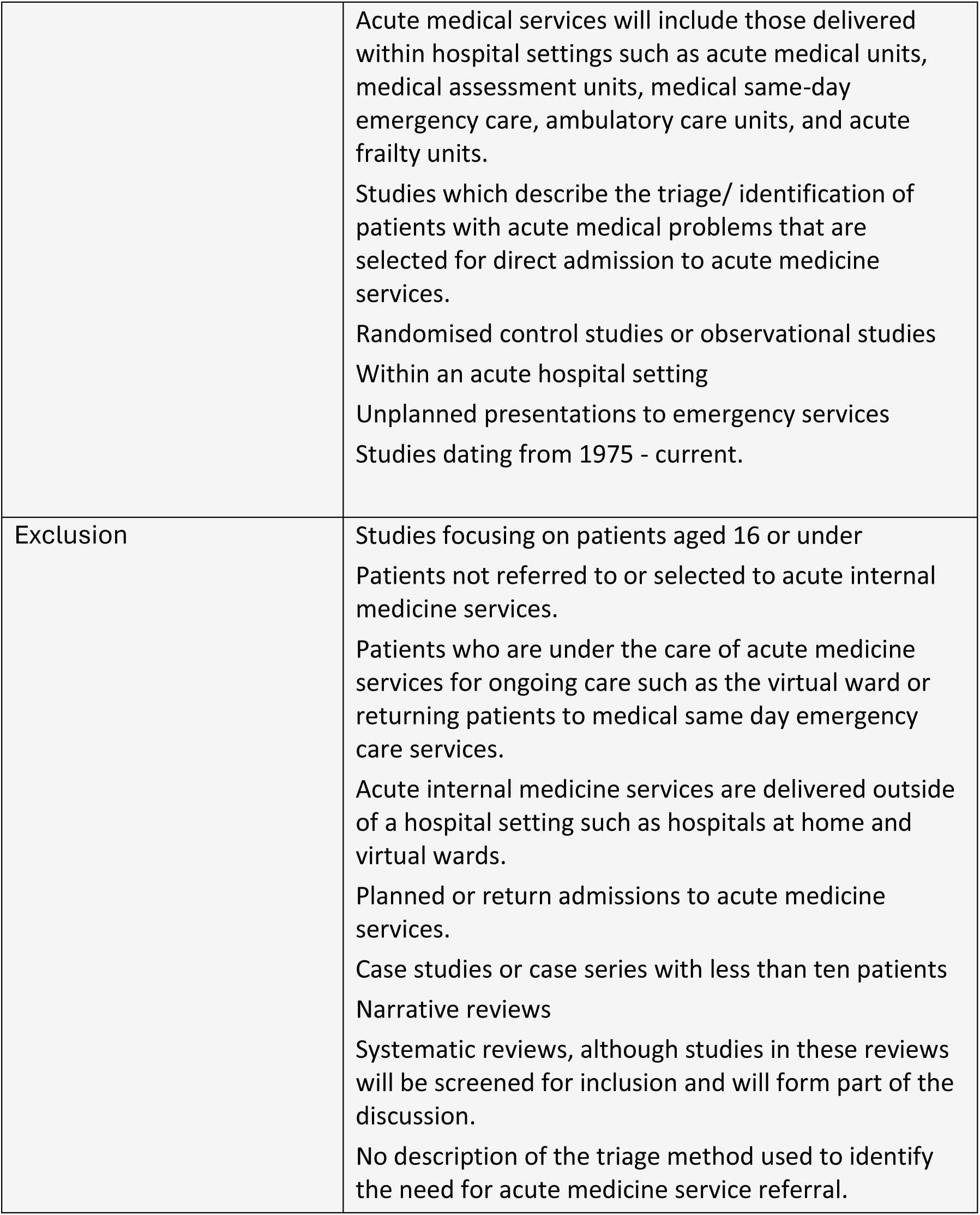
Inclusion and Exclusion Criteria for studies to be included in the Systematic Review.

|  |  |
| --- | --- |
| Inclusion | Adult Patients (over 16 years of age) presenting to the emergency departments who require acute medical assessment. |
|  | Acute medical services will include those delivered within hospital settings such as acute medical units, medical assessment units, medical same-day emergency care, ambulatory care units, and acute frailty units.<br>Studies which describe the triage/ identification of patients with acute medical problems that are selected for direct admission to acute medicine services.<br>Randomised control studies or observational studies Within an acute hospital setting<br>Unplanned presentations to emergency services<br>Studies dating from 1975 - current. |
| Exclusion | Studies focusing on patients aged 16 or under<br>Patients not referred to or selected to acute internal medicine services.<br>Patients who are under the care of acute medicine services for ongoing care such as the virtual ward or returning patients to medical same day emergency care services.<br>Acute internal medicine services are delivered outside of a hospital setting such as hospitals at home and virtual wards.<br>Planned or return admissions to acute medicine services.<br>Case studies or case series with less than ten patients<br>Narrative reviews<br>Systematic reviews, although studies in these reviews will be screened for inclusion and will form part of the discussion.<br>No description of the triage method used to identify the need for acute medicine service referral. |

### Data extraction & quality assessment

Data were extracted from eligible full texts onto a pre-designed form by two independent reviewers (see supplementary materials). This included data relating to any of the review outcomes above and descriptions of the model used to select patients for direct medical assessment pathways. Where data was not reported, study authors were contacted a maximum of three times to request data.

### Risk of bias assessment

Studies were assessed for possible bias using the ROBINS-I tool^[9]^. This considers both bias and applicability to the review question across key domains of the intervention (here, the direct admission pathway), including its definition and application; patient selection; potential confounding influences; and conduction and interpretation of the results.

### Data synthesis

The heterogeneity of included studies prevented a meta-analysis and so a narrative synthesis was performed. Subgroup analysis for each outcome was performed based on the tool used to identify patients for direct admission services and the outcomes reported by the studies.

## Results

The selection process is shown in the PRISMA flow diagram (Figure 1) including reasons for exclusion. The search found 6878 articles. Once duplicates were removed (2473 in total), 89 articles were eligible for full text screening. From these, four articles were included for data extraction (Table 3).

**Figure 1:**
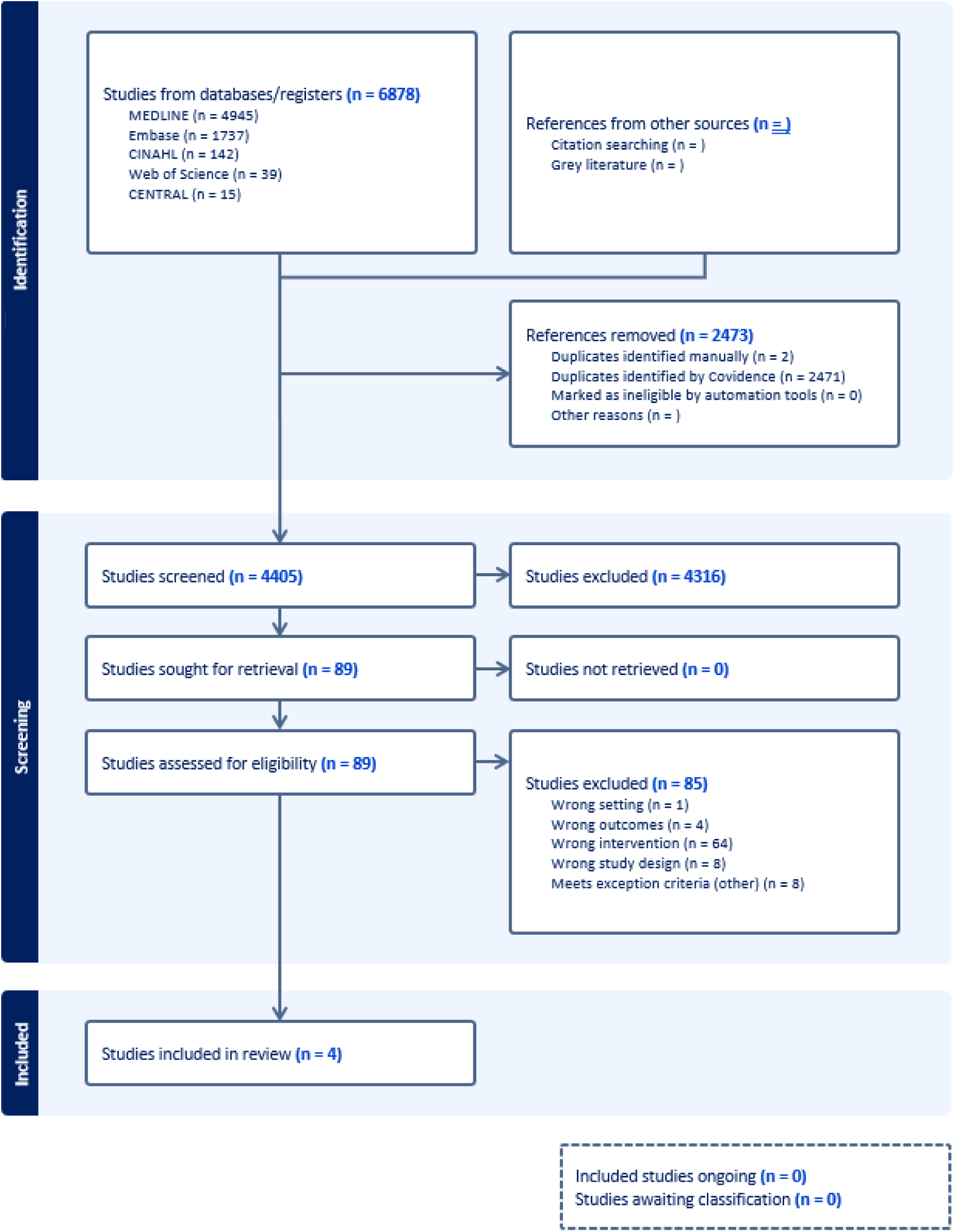
**PRISMA Flow diagram**

**Table 3:** Studies included for final extraction.

| Author | Year | Type of Study | Country | Number of participants | Total participants | Inclusion criteria | Exclusion criteria | Primary Outcome | Secondary outcome | Patient Selection tool | Comparator | Results |
| --- | --- | --- | --- | --- | --- | --- | --- | --- | --- | --- | --- | --- |
| R. SCHOENE<br>NBERGER<br>[10] | 1992 | Prospective observational, single centre | Switzerland | 974 | 2431 emergency department patients | medical inpatients who had been transferred from the emergency room to general internal medicine wards. Data collected from Feb 1990 – April 1990. | Patients who had died during the index in-hospital stay | Assessment of initial triage decisions by actual outcomes after hospital admission | Not specified | Internal Medicine resident triage following referral to medicine from ED into 4 categories:<br>A. Acutely ill patients needing care on an acute medical ward for undetermined durations but longer than two to three days.<br>B. Acutely ill patients needing care on an acute medical ward who's in hospital stays will not exceed two to three days.<br>C. Chronically ill and disabled patients with realistic prospects for rehabilitation leading to a return to the previous social setting.<br>D. Chronically ill and disabled patients who will stay personally dependent and who will need institutionalization in a skilled nursing care facility. | Nil | Standard formulas were used to calculate sensitivity, specificity, predictive values, and accuracies of the initial triage decision. Counts were compared with chi-square testing, continuous variables with Student's t-test. A two-tailed p-value < 0.05 was considered significant.<br>Initial triage was correct 95% of the time for acutely unwell patients (category A and B).<br>Patients defined as chronically ill (C and D) the initial triage was correct 67% of the time with 56% of those in category D being rehabilitated. |
| A. Emmanuel<sup>[11]</sup> | 2010 | Retrospective observational, single centre | Ireland | 270 | 270 emergency department patients | All medical admissions aged 14 years or older. Data collected from April 2009 – June 2009. | Patients who were not admitted to hospital | The use of CTS and SCS to predict the need for hospital admission through risk of mortality | Not specified | Cape Triage Score | Nil | The ability of CTS to predict in-hospital mortality was statistically inferior to the SCS. AUROC for in hospital death was 0.94 for the SCS and 0.64 for the CTS. |
| M. Zahid<sup>[12]</sup> | 2023 | Retrospective cross-sectional, single centre | Qatar | 320 299 | 370217 emergency department patients | Patients > 14 years of age who presented to the ED with medical complaints and were evaluated with vital signs measurement during the study period were included. Data collected from Jan 2019 – Dec 2019. | Patients who left the ED before a decision of admission could be made, those presenting with non-medical complaints (trauma, fractures, gynaecological, acute surgical, psychiatric, Ophthalmologic, and otorhinolaryngologic complaints), those who did not have vital signs recorded and who died in the ED before a decision to admit could be made, were excluded. | The ability of the medical admission prediction score to predict medical admission | Not specified | Medical Admission Prediction Score | Nil | The model had a sensitivity of 69.1% (95% CI 68.2–69.9), specificity was 83.9% (95% CI 83.7–84.0), positive predictive value (PPV) 14.2% (95% CI 13.8–14.4), negative predictive value (NPV) 98.6% (95% CI 98.5–98.7) and positive likelihood ratio (LR+) 4.28% (95% CI 4.27–4.28). |
| C. Bartlett<br>[13] | 2023 | Prospective observational, single centre | USA | 74 | 74 emergency department patients | ED admission requests to internal medicine. Data collected from a pilot period from Nov 2019 – Dec 2019 | Not specified | Time to admission (TTA)—the time between an ED admission request and internal medicine (IM) admission orders | ICU transfers, Length of Stay, ED crowding, interprofessional practice attitudes between IM and ED clinicians. | Internal Medicine resident triage following referral to medicine from ED | Nil | TTA 5 hours 19 minutes (median, 4 hours 45 minutes) to a postintervention average of 2 hours 8 min.<br>ED-2 as an indicator of crowding in ED demonstrated a statistically significant upward shift, interpretation of this is complicated by the COVID19 pandemic during the study period.<br>ICU admissions 1.1% pre intervention to 1.4% post intervention<br>Length of stay 6.48 days vs 6.62 days.<br>Interprofessional practice perceptions improved in the EM team but not the IM team. |

Abbreviations: Cape Triage Score (CTS), Simple Clinical Score (SCS), Time to Admission (TTA), Internal Medicine (IM), ED-2 (median time from admit decision to departure from ED for inpatient admission), Intensive Care Unit (ICU), Emergency Department (ED), Confidence Interval (CI), Area Under Receiver Operating Characteristic Curve (AUROC), Positive Predictive Value (PPV), Negative Predictive Value (NPV), Positive Likelihood Ratio (LR+), Medical Admission Score (MAP).

All studies were observational and two were retrospective. The countries of which each study was undertaken varied significantly and shows a broad geographical distribution (Qatar, Ireland, USA and Switzerland), and all were performed in single centres. The variance in publication date was also notable with one study being 32 years since publication and the other over 10 years. The remaining two studies were published in the past year.

In total 323,074 individual patients were included in all studies combined. A significant amount of the sample was provided by one single study with the remaining studies having sample sizes ranging from 2431 to 74. The largest study (320,299 sample size) had a predominantly Asian (53.7%) and Arabic demographic (38.7%). Appropriate age data could only be collected for 3 of the studies with a mean age of 62.7. The risk of bias of the 4 included studies can be found in Figure 2.

**Figure 2:**
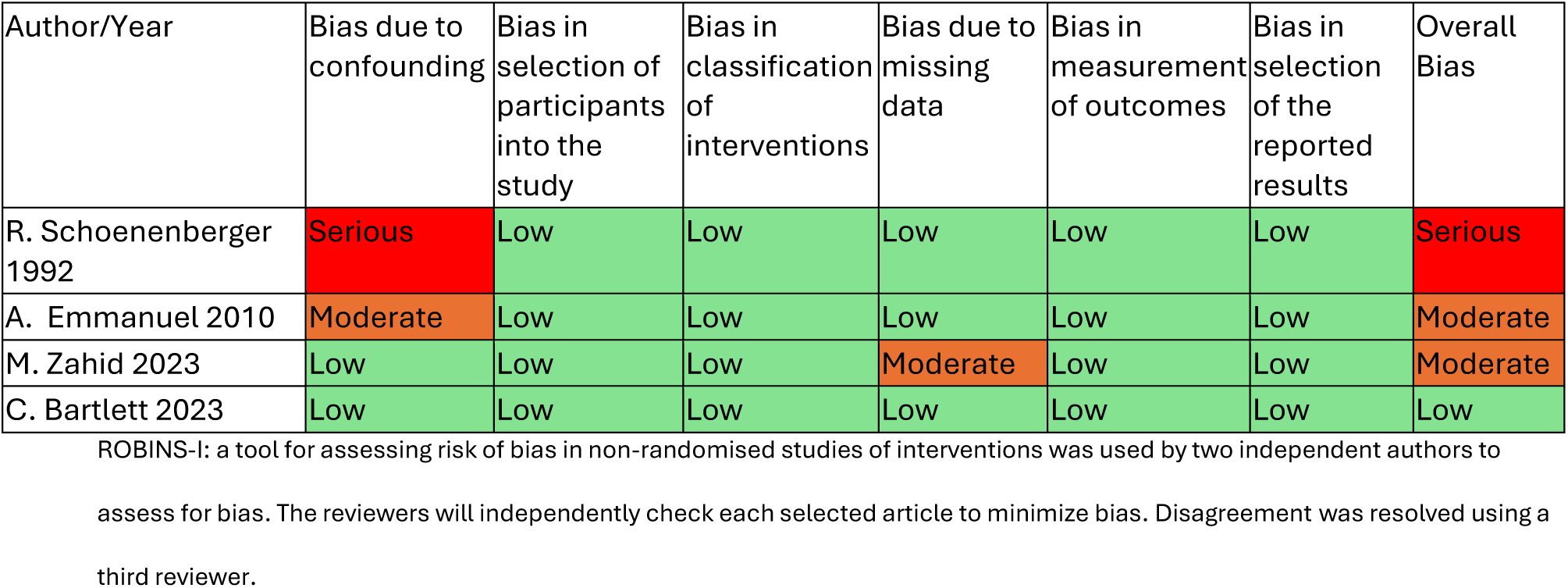
**Risk of Bias Assessment**

The four studies showed a variable risk of bias. Schoenenberger et al^[10]^ had a significant risk of bias due to confounding factors. The study did not provide information on the standardised admission and examination proforma used and how the patients were allocated to categories A-D. Emmanuel et al^[11]^ was found to have moderate risk of bias due to confounding as health demographics were not recorded and therefore these variables and co-morbidities could have an influencing effect on the performance of the CTS and SCS which were not included in the analysis. Zahid^[12]^ was deemed to have a moderate risk of bias due to missing data of patients who did not have observations recorded. Bartlett et al ^[13]^ was found to have a low risk of bias across all domains.

### Results of included studies

Of the four articles, Emmanuel 2010^[11]^ & Zahid 2023^[12]^ focused on the development of an admission prediction tool to assess the likelihood of medical admission or mortality.

Emmanuel et al^[11]^ conducted a prospective observational study in Ireland where they developed a Cape Triage Score (CTS) to be used alongside the pre-existing Simple Clinical Score (SCS) that was already in routine use. The study examined 2094 attendances to the emergency department between (April 2009 – June 2009) of which there were 270 admissions under medicine which were included in the study. The study did not include a direct admission pathway to acute medicine and all patients were seen through the traditional pathway by an emergency physician. The article found that the Cape Triage Scores ability to predict in-hospital mortality was statistically inferior to the simple clinical score, AUROC 0.64 vs 0.94 (p<0.002). However, the Cape Triage Score was reported to be more effective at identifying medical admissions in cohorts that would traditionally score low in the SCS (such as dizziness, syncope and palpitations) and therefore indicated a potential for the SCS to not capture these patients that required admission.

Zahid et al^[12]^ conducted a retrospective cross-sectional analysis of 320,229 medical patients admitted to Hamad General Hospital in Qatar between (January 2019 – December 2019). The included patients were >14 years of age presenting to the emergency department with a medical complaint. Patients that were excluded were those who self-discharged, had non-medical presentations and any without vital signs recorded. The aim was to develop a scoring system predicting hospital admission to a medical ward for patients presenting in ED from 9 variables which they called the medical admission prediction score (MAPS). These included two demographic variables (age and sex) and seven clinical variables (pulse rate, respiratory rate, systolic blood pressure, oxygen saturation, Glasgow coma scale, number of comorbidities, and hospitalization in the last 30 days). The primary objective of this study was to develop a tool which could be used to predict admission risk that did not rely upon other triage tools such as the Manchester Triage Score (MTS) which other admission prediction tools use. The authors put forward that the MAPS is more versatile than other international admission prediction scores (such as the Simple Triage And Rapid Treatment (START) and Glasgow Admission Prediction Score (GAPS)) due to this The MAPS was shown to have a higher odds ratio for admission, 2.68 (95%CI 2.53-2.84, p< 0.001) than START 1.93 (95%CI 1.90-1.96, p< 0.001). However, in a sub-analysis MAPS odds ratio decreased in patients who were >60 years to 1.88 (95% CI 1.77, 2.01, p< 0.01) compared to START which increases to 3.31 (95% CI 3.25, 3.37) for 60–79-year-old and to 6.01 (95% CI 5.89, 6.13) for > 80 years old patients. The authors performed a Receiver Operator Characteristic (ROC) analysis comparing admission to medicine and chose a cut off value of >17. The area under the curve (AUC) was 0.831 (95% CI 0.827– 0.836) with a predictive accuracy of 83.3% (95% CI 83.2–83.4). The sensitivity at this cut-off was 69.1% (95% CI 68.2–69.9) and specificity was 83.9% (95%CI 83.7–84.0). The positive predictive value was 14.2% (95% CI 13.8–14.4), negative predictive value 98.6% (95% CI 98.5–98.7) and positive likelihood ratio 4.28% (95% CI 4.27–4.28).

The remaining two articles (Schoenenberger et al^[10]^ & Bartlett et al^[13]^) assessed the use of a medical triage clinician in the emergency department.

Schoenenberger et al^[10]^ conducted a prospective observational study in Switzerland of 2431 emergency department attending patients between February 1990 – April 1990, investigating the direct triage of emergency department patients by a senior resident doctor and designated into 4 distinct categories (included in table 3) based on acuity, expected duration of care and rehabilitation potential. This was in addition to the traditional pathway of assessment in the emergency department who deemed the patient as requiring an internal medicine admission. There was no comparator group in the study to reflect the traditional route of admission. The assessment conducted by the resident doctor involved the use of a standardised admission sheet and examination combined with clinical judgement. Primary outcome measured was the accuracy of initial triage by actual outcomes of index hospitalizations. Initial triage was correct 95% of the time for acutely unwell patients (category A and B). For patients defined as chronically ill (C and D) the initial triage was correct 67% of the time with 56% of those in category D being rehabilitated, showing a PPV of 44%, with the triage assessment overestimating the need for a long-term nursing care facility. The study concluded that clinical judgement in the emergency department by internal medicine specialists can distinguish acutely unwell from chronically unwell patients.

Bartlett et al^[13]^ conduced a prospective observational study of 74 emergency department patients with a pilot period from November 2019 – December 2019, who were triaged in the emergency department following an internal medicine admission request by the emergency department team over the course of eight pilot shifts. The primary outcome measure was the time to admission (TTA). The comparator population were patients referred to IM in the pre-intervention Period. Participants were selected based on clinical assessment by the emergency department team with the criteria for inclusion being that they required and internal medicine admission. Exclusion criteria patients that were not referred for an IM inpatient bed and were not included in the study. The findings were that time to admission decreased following this intervention from 5 hours 19 minutes (median 4 hours 45 minutes) to 2 hours 8 minutes. This intervention did not change length of stay (6.48 days vs 6.62 days). There was no change in admissions to ICU (1.1% pre intervention to 1.4% post intervention admissions). ED-2 (median time elapsed from admit decision time to time of departure from the ED for patients admitted to inpatient status) was used as an indicator of crowding in ED, and demonstrated a statistically significant upward shift, with more ED overcrowding, however, the authors suggested that interpretation of this was complicated by the COVID19 pandemic during the study period. The authors also conducted interviews with medical staff (ED and IM resident, attending and advance practice providers).122 of 309 (preintervention group) and 98 of the 309 (post intervention group) responded. The results showed that ED residents and attendings demonstrated a statistically significant improvement in interprofessional practice perspectives with IM. In the post intervention ED group it showed statistically significant worsening in the perception that processed to admitting patients to a internal medicine service is difficult. The author put forwards that this may be due to 25% of referrals being identified by the IM triage for an alternative destination.

## Discussion

This systematic review has assessed the evidence to support direct admission pathways from urgent and emergency care triage to acute medical assessment, excluding the traditional steps of assessment by Emergency medical staff.

The four studies included following an extensive literature search were heterogenous and of variable quality. Only one study was based in a health system comparable to the National Health Service in the UK, with this study conducted over 14 years ago, arguably when pressures on acute services were less than they are now.

Two studies aimed to develop an admission prediction tool to assess the likelihood of medical admission or mortality. The admission prediction scores examined in these papers attempt to capture the acuity of the patient’s illness by utilising vital signs and other risk factors for severity of illness and risk of mortality. The performance of these scores across different subgroups of patients varied, and neither score was used to directly stream patients to medicine or has been validated as a tool for direct admission to medicine in further randomised controlled studies or in a different healthcare setting. The CTS goes some way to begin defining some clinical presentations that do not overtly score highly on vital sign monitoring, such as a patient presenting with chest pain who may need medical admission ^[11]^. Whilst these scores do capture to some extent the acuity of illness and risk of in hospital mortality, Emmanuel 2010 ^[11]^ demonstrated that many of these scores were not able to accurately identify medical patients that will require a hospital admission. There is currently insufficient evidence to support the use of either tool in a direct admission pathway.

The other two studies assessed the use of a triage clinician with internal medicine training in the emergency department. Bartlett et al ^[13]^ demonstrated that having the triage clinician be a point of contact for medical referrals through the emergency department improves TTA however, this was not a direct streaming pathway to acute medicine and all patients still had a full assessment by emergency department clinicians prior to being referred. The study reports it was impacted by COVID19 pandemic. This impact, alongside the small study size and the single centre setting limits the interpretation of results but does demonstrate a potential time benefit for patients to be seen by the most relevant clinical team (here, medicine) compared to standardised pathways.

Schoenenberger et al ^[10]^ utilised the triage clinician in an alternative way, which was more reflective of a direct medical assessment in the emergency department although all patients had an emergency department assessment prior to the IM triage and the focus of the triage clinician was to consider acuity and rehabilitation potential. The study assessed the direct triage of these patients by a senior medical clinician and their ability to predict acuity, length of stay and rehabilitation potential. However, the study was conducted in a time which is unlikely to reflect modern challenges faced by urgent and emergency services. The study does indicate the potential value for early clinical review by an internal medicine specialist to triage patients to acute medicine services.

Overall, there remains a lack of evidence for pathways which stream patients from triage directly to medicine, despite clear theoretical gains from avoiding duplication in assessments and enabling patients to be seen by the most appropriate medical team as quicky as possible. It is unclear how these pathways should be configured, how they should be staffed, wo should be triaging to medicine and what tools should be used to select patients. Future research should focus on building the evidence base for a direct admission pathway that would standardise and reduce the variation currently seen across acute medicine services. A limitation within this systematic review is that there is significant variation in terminology used when referring to these acute medical services internationally, although terms used in searches were broadened to specifically accommodate this.

## Conclusion

There is a critical need to change urgent and acute care pathways to improve patient outcomes and experience and reduce pressures on emergency and acute medical teams. There is significant interest in direct care pathways but this systematic review highlights there is currently little published evidence to guide how they should be developed with no studies addressing this question directly. Further studies are needed to assess existing pathways and to implement any changes to improve patient care and flow.

## Supporting information

Record of Database Searches

Risk of Bias Assessment Table

## Data Statement

Any data that has not been included in the manuscript will be available on request or will be included in supplementary documents.

## Data Availability

All data produced in the present study are available upon reasonable request to the authors

