## Supplementary material for "What strategies are used to select patients for direct admission under acute medicine services? A systematic review of the literature": Record of Database Searches

Example Search Strategy:

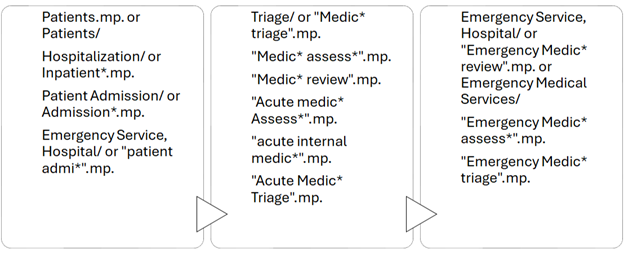

Record of Database Searches

**CENTRAL (15)**

Search Name: CENTRAL

Date Run: 17/01/2024 20:14:52

ID Search Hits

#1	(patient) OR (Inpatient*) OR (Admission*) OR (Patient NEXT admi*) (Word variations have been searched)	1231187

#2	(Medic* NEXT triage) OR (Medic* NEXT assess*) OR (Medic* NEXT review) OR (Acute NEXT medic* NEXT Assess*) AND (acute NEXT internal NEXT medic*) (Word variations have been searched)	3285

#3	(Emergency NEXT Medic* NEXT review) OR (Emergency NEXT Medic NEXT assess*) OR (Emergency NEXT Medic* NEXT triage) OR (Emergency NEXT medical NEXT services) (Word variations have been searched)	1970

#4	#1 AND #2 AND #3	15

**CINAHL (143)**

\# Query Limiters/Expanders Last Run Via Results S4 S1 AND S2 AND S3 Limiters - Publication Year: 1975-2023 Expanders - Apply equivalent subjects Narrow by SubjectAge: - all adult. Search modes - Boolean/Phrase Interface - EBSCOhost Research Databases Search Screen - Advanced Search Database - CINAHL Plus 143 S3 TX “Emergency Medic* review” OR TX “Emergency Medic* assess*” OR TX “Emergency Medic* triage” OR TX emergency medical services OR TX emergency department Expanders - Apply equivalent subjects Search modes - Boolean/Phrase Interface - EBSCOhost Research Databases Search Screen - Advanced Search Database - CINAHL Plus 170,567 S2 TX “Medic* triage” OR TX “Medic* assess*” OR TX “Medic* review” OR TX “Acute medic* Assess*” OR TX acute internal medicine OR TX “Acute Medic* Triage” Expanders - Apply equivalent subjects Search modes - Boolean/Phrase Interface - EBSCOhost Research Databases Search Screen - Advanced Search Database - CINAHL Plus 5,720 S1 TX patients in hospital OR TX ( inpatient or acute or hospital or ward or unit ) OR TX admission to hospital OR TX patient admission Expanders - Apply equivalent subjects Search modes - Boolean/Phrase Interface - EBSCOhost Research Databases Search Screen - Advanced Search Database - CINAHL Plus 2,606,116

**EMBASE (1737)**

1	Patients.mp. or patient/	10992542

2	hospital patient/ or Inpatient*.mp.	348330

3	hospital admission/ or Admission*.mp.	667317

4	"Patient admi*".mp.	11112

5	1 or 2 or 3 or 4	11247071

6	"Medic* triage".mp.	179

7	medical assessment/ or "Medic* assess*".mp.	38547

8	"Medic* review".mp.	6490

9	"Acute medic* Assess*".mp.	112

10	internal medicine/ or "acute internal medic*".mp.	46528

11	"Acute Medic* Triage".mp.	2

12	6 or 7 or 8 or 9 or 10 or 11	91449

13	"Emergency Medic* review".mp.	25

14	"Emergency Medic* assess*".mp.	21

15	emergency health service/ or "Emergency Medic* triage".mp. or emergency ward/	322234

16	13 or 14 or 15	322261

17	5 and 12 and 16	3792

18	limit 17 to (human and yr="1975 -Current")	3582

19	limit 18 to (adult <18 to 64 years> or aged <65+ years>)	1737

**Medline ALL (2570)**

Ovid MEDLINE(R) ALL <1946 to January 16, 2024>

1	Patients.mp. or Patients/	7310276

2	Hospitalization/ or Inpatient*.mp.	270807

3	Patient Admission/ or Admission*.mp.	309083

4	Emergency Service, Hospital/ or "patient admi*".mp.	116413

5	1 or 2 or 3 or 4	7511383

6	Triage/ or "Medic* triage".mp.	15311

7	"Medic* assess*".mp.	2724

8	"Medic* review".mp.	3303

9	"Acute medic* Assess*".mp.	45

10	"acute internal medic*".mp.	69

11	"Acute Medic* Triage".mp.	2

12	6 or 7 or 8 or 9 or 10 or 11	21321

13	Emergency Service, Hospital/ or "Emergency Medic* review".mp. or Emergency Medical Services/	134094

14	"Emergency Medic* assess*".mp.	11

15	"Emergency Medic* triage".mp.	20

16	13 or 14 or 15	134110

17	5 and 12 and 16	6187

18	limit 17 to (humans and yr="1975 -Current")	5998

19	limit 18 to "all adult (19 plus years)"	2570

**Medline in process (2375)**

Ovid MEDLINE(R) and In-Process, In-Data-Review & Other Non-Indexed Citations <1946 to January 23, 2024>

1	Patients.mp. or Patients/	7218005

2	Inpatients/ or Hospitalization/ or Inpatient*.mp.	267881

3	Patient Admission/ or Admission*.mp.	304536

4	"Patient admi*".mp.	31365

5	1 or 2 or 3 or 4	7382668

6	Triage/ or "Medic* triage".mp.	15291

7	"Medic* assess*".mp.	2684

8	"Medic* review".mp.	3246

9	"Acute medic* Assess*".mp.	45

10	"acute internal medic*".mp.	68

11	"Acute Medic* Triage".mp.	2

12	6 or 7 or 8 or 9 or 10 or 11	21203

13	Emergency Service, Hospital/ or "Emergency Medic* review".mp. or Emergency Medical Services/	133984

14	"Emergency Medic* assess*".mp.	11

15	"Emergency Medic* triage".mp.	20

16	13 or 14 or 15	134000

17	5 and 12 and 16	4900

18	limit 17 to humans	4812

19	limit 18 to "all adult (19 plus years)"	2375

**Web of Science (39)**

Web of Science Search Strategy (v0.1)

Search: (((ALL=(Patients)) OR ALL=(inpatient*)) OR ALL=(Admission*)) OR ALL=("patient

admi*") Editions: WOS.SCI,WOS.SSCI,WOS.ISTP,WOS.ISSHP,WOS.ESCI Date Run: Thu

Apr 18 2024 08:42:16 GMT+0100 (British Summer Time) Results: 8610302

Search: ((ALL=(“Emergency Medic* assess*”)) OR ALL=(“Emergency Medic* review”)) OR

ALL=(“Emergency Medic* Triage”) Editions:

WOS.SCI,WOS.SSCI,WOS.ISTP,WOS.ISSHP,WOS.ESCI Date Run: Tue Jan 23 2024

08:42:16 GMT+0100 (British Summer Time) Results: 57

Search: (((((ALL=("medic* triage")) OR ALL=("medic* assess*")) OR ALL=("medic* review")) OR

ALL=("acute medic* assess*")) OR ALL=("acute medic* triage")) OR ALL=(“acute internal

medic*”) Editions: WOS.SCI,WOS.SSCI,WOS.ISTP,WOS.ISSHP,WOS.ESCI Date Run: Thu

Tue Jan 23 2024 08:42:17 GMT+0100 (British Summer Time) Results: 8016

\# Database: Web of Science Core Collection

\# Entitlements:

- WOS.IC: 1993 to 2024

- WOS.CCR: 1985 to 2024

- WOS.SCI: 1900 to 2024

- WOS.AHCI: 1975 to 2024

- WOS.BHCI: 2010 to 2024

- WOS.BSCI: 2010 to 2024

- WOS.ESCI: 2005 to 2024

- WOS.ISTP: 1990 to 2024

- WOS.SSCI: 1900 to 2024

- WOS.ISSHP: 1990 to 2024

\# Searches:

Search: #1 AND #2 AND #3 Editions:

WOS.SCI,WOS.SSCI,WOS.ISTP,WOS.ISSHP,WOS.ESCI Timespan: 1975-01-01 to

2024-01-17 Date Run: Tue Jan 23 2024 08:42:17 GMT+0100 (British Summer Time) Results: 39
